# Preventive psychosocial services and collaboration for children and families: protocol for a mixed-methods intersectoral mapping study at community level

**DOI:** 10.64898/2026.05.27.26354209

**Authors:** A. Reinhart, S. Beierle, L. Popp, B. Voigt, S. Schneider, B. Reißig, S. Walper, S. Kuger, A. Alayli, F. De Bock

**Affiliations:** Charité – Universitätsmedizin Berlin, corporate member of Freie Universität Berlin and Humboldt-Universität zu Berlin, Institut für Medizinische Soziologie und Rehabilitationswissenschaft, Berlin, Germany; German Youth Institute (DJI), Munich / Halle (Saale), Germany; Mental Health Research and Treatment Center (FBZ), Ruhr University Bochum, Bochum, Germany; Ludwig Maximilians University, Faculty of Psychology and Education, Munich, Germany; Child Health Services Research Unit, Department for General Pediatrics, Neonatology and Pediatric Cardiology, Medical Faculty and University Hospital Düsseldorf, Heinrich Heine University Düsseldorf, Düsseldorf, Germany; German Center for Mental Health (DZPG), Bochum-Marburg site, Bochum, Germany

**Author notes:** Corresponding Author: Reinhart, A. Anika Reinhart. Email addresses for each author: Sarah Beierle, Lukka Popp, Babett Voigt, Silvia Schneider, Birgit Reißig, Sabine Walper, Susanne Kuger, Adrienne Alayli, Freia De Bock.

**Keywords:** psychosocial services, mental health, primary prevention, secondary prevention, support system, intersectoral collaboration, children, families, service mapping, network

## Abstract

**Background:** Many mental health problems originate in childhood, highlighting the need for early preventive approaches. Preventive services to promote children’s mental health are offered in the health, education, and social sectors (H-E-S) but are often not used by certain at-risk groups or early enough. To identify children at-risk and provide needs-oriented support, professionals from all sectors must be well trained, collaborate closely to refer to specialized services for specific mental health problems or risk factors, and understand the regional psychosocial support system and its services. A comprehensive approach to preventing mental health problems requires structured planning and a systematic overview of all institutions and services in the region and their collaboration. This study aims to map the preventive mental health and psychosocial support service^1^ system and the collaboration between institutions across three sectors (H-E-S) in two exemplary city districts. The study is integrated into a whole-district approach to child mental health promotion that is being implemented in one of the researched city districts, and its results will inform further activities there.

**Methods:** We use a mixed-methods approach, combining qualitative interviews with a quantitative survey to map psychosocial services for children aged 4 to 10 and their families across the H-E-S sectors in two socioeconomically disadvantaged city districts in East and West Germany. All institutions that potentially offer psychosocial services for children and families will be approached to recruit professionals (e.g., schools, practices, counseling centers).

To understand the regional psychosocial support system, we will analyze existing services and their characteristics (e.g., target groups, intervention types) descriptively. Social network analysis will be applied to gain an in-depth understanding of collaboration between institutions, to identify potential gaps in services and pathways, and to inform an intervention aimed at improving interinstitutional and intersectoral collaboration.

**Discussion:** To our knowledge, this is the first study to comprehensively analyze regional preventive psychosocial support systems for children and families across sectors at the community level. Previous mappings of psychosocial services have focused on a single sector (e.g., health) or specific diagnoses only. The psychosocial preventive landscape spanning the H-E-S sectors involves complex financing structures and referral logics. Understanding the characteristics of the existing support landscape requires a systematic and comprehensive approach. Our study advances service mapping and operationalization methods in public health research. Additionally, the findings will inform recommendations for improving comprehensive prevention approaches in the selected city districts.

**Ethics and dissemination:** Ethical approval has been obtained. Study findings will be disseminated through publications and conference presentations.

**Trial registration:** no

**Article summary:** *Strengths and limitations of this study:*

- First scientific attempt to map the psychosocial prevention landscape for children and families at city district level, comprising services and institutions across the health, education, and social sectors (strength)
- Understanding how psychosocial prevention works at city district level (strength)
- Starting point for advancing structural-level approaches (e.g., strengthening intersectoral collaboration) (strength)
- Focus on city districts; other spatial units (e.g. rural areas) have not yet been considered (limitation)
- The survey focuses on professionals, the client perspective is not yet included (limitation)

## 1. Background

### Mental health problems in childhood and early detection of risk factors

Mental health disorders are among the most common conditions for children and young people worldwide, with a point prevalence of at least 15% and a lifetime prevalence of up to one third until adulthood (1). In addition, up to 20% of children and young people have mental health problems, e.g., emotional or behavioral issues like sadness, aggression, insomnia, difficulties in school, family and/or social environments (2). Early mental health problems predispose to the development of mental health disorders and, even if thresholds for an explicit mental health disorder are not exceeded, can be associated with unfavourable developmental trajectories (3).

In childhood and adolescence, mental health problems are often caused by psychosocial risk factors in families, such as parental mental health problems or addiction, the experience of violence, poverty, or a lack of social support (4). These factors, especially if combined, can undermine parenting skills and increase child abuse or neglect (5, 6). Exposure to these adverse conditions across the early years is a key predictor of mental health problems (7).

In order to prevent mental health problems in childhood and to avoid further worsening, it is crucial to identify psychosocial risk factors and mental health problems at an early stage and to provide psychosocial support targeted to individual or family needs (7), especially given the recent collective experience of social disruption during the pandemic. If implemented early, targeted psychosocial support has been shown to promote mental well-being and create positive mental health prospects in the long term (3, 8–10).

Professionals from various disciplines, institutions, and sectors, i.e., health, education, and social sectors (H-E-S), are relevant for the early identification of psychosocial risk factors and mental health problems, as well as the provision of needs-oriented psychosocial support. Professionals that have frequent contact with children and parents, such as early childcare and schoolteachers, or social workers can detect changes in mood or behavior in children, refer to suitable services or provide such services themselves (11–14).

### National and international research findings on early detection and collaboration

Institutions, professionals, and psychosocial services together form a landscape of psychosocial support with the potential to act as a system to prevent mental health problems for children (15, 16). However, many psychosocial services are not fully used, not used early enough, or not used by certain at-risk groups (17, 18). International research suggests that one underlying reason for the lack of psychosocial service use is that professionals are often unaware of the services available in their local area, particularly those provided outside their own sector (19, 20, 17). But there are also services that are difficult to access quickly due to limited capacity and long waiting lists (i.e., medical services). A functioning psychosocial support system should not only enhance the ability of professionals to act as gatekeepers and multipliers within the system but also tackle barriers to effective use so that children and families with psychosocial support needs receive needs-oriented psychosocial support at an early stage. Accordingly, the World Health Organization (WHO) sees intersectoral collaboration between professionals from different sectors as the key to preventing mental health problems (21). Research indicates that successful collaboration between professionals within and between H-E-S sectors can help clients accessing psychosocial services tailored to the individual needs and prevent underserved populations from being excluded from support (21–24).

### Mapping research

To facilitate more effective prevention, a comprehensive and systematized prevention approach is required that considers the entire regional psychosocial service landscape. This implies a population health perspective to the prevention of mental health problems, considering the region as a support system providing service functions and structures within various sectors for a regionally determined child population (25, 26). The population members who use or are intended to use this service landscape as clients are children and their parents. An intersectoral system perspective enables understanding the functioning and structure of the support system with its various sectors from a systems’ perspective to develop approaches to improve prevention (16).

To support regional planning from a population health perspective, several service mapping tools have been developed, internationally. Seemingly well-established tools, yet on a country or macro-level, are, for example, the European Service Mapping Schedule (ESMS), the subsequent Description and Evaluation of Services and Directories for Long Term Care (DESDE-LTC), and European Child and the Adolescent Mental Health Mapping Questionnaire (ECM-Q) (27–30). Mapping allows analyzing a) which psychosocial services are available or lacking in certain regions; b) which actors and institutions are involved in the psychosocial support system of children and families; and c) which actors and services should be part of the support system, but currently are missing (27, 31). These existing mapping tools provide categorization criteria for identified services. For example, psychiatric care services are categorized according to type/form of service, location or accessibility, provider, availability, target group, funding, function of the service, or structure of the facility offering the service (27). The available mapping tools are subject to several limitations, however. They only map services on a higher level (i.e., the region or country, not community level), only map for adults (not children), and/or only map for psychiatric health care (not prevention) (32). Additionally, they rarely consider services outside the health sector and applications in Germany are still rare (27–29).

### Existing Research on the Psychosocial Support System in Germany

Research on the German psychosocial support system to promote mental health primarily focuses on services for expectant parents and families with children up to age 3. These services, called Early Childhood Intervention (“Frühe Hilfen”) in Germany, include screening for psychosocial needs in maternity wards, and home visitation programs for young families to low-barrier parent-child centers in Germany. The program “Frühe Hilfen” is implemented at the community level but financed by a federal foundation. Special attention has been paid to the establishment of services and intersectoral collaboration between the healthcare system and youth welfare services. In 2020, there were 939 early help networks in 570 municipalities in Germany (33). While this support system is well established for families with very young children, families with children over the age of three, i.e., at pre-school and primary school age, have received less attention.

Some municipalities in Germany have established a “prevention chain” (Präventionskette) model of care for children from highly burdened families. Prevention chains are community strategies that combine different services and focus on important transitions (e.g., transition from kindergarten to school) for specific vulnerable groups of children (34). The aim is to prevent vulnerable children from being left behind without help, e.g., during the transition from family to preschool and preschool to school age. However, the prevention chain approach only exists in certain selected communities, does not involve health services well enough, is not well researched, not described in the public health literature, and lacks evidence on its effectiveness and functions (35).

### Objectives

This study aims to analyze and understand the psychosocial support system and its services for children and families at the community level by conducting a mapping of the existing landscape of primary and secondary preventive psychosocial services and the collaboration between institutions and professionals across the H-E-S sectors in two socioeconomically disadvantaged city districts within two major German cities.

We assume that the characteristics of the preventive psychosocial support services for children and families will differ from those of previous mapping studies (e.g., in psychiatric care), and that the target group (children, families) will make a difference in respective services (36, 37). Therefore, existing mapping procedures and service categorization systems can only be used to a limited extent, but rather new categories and survey instruments must be developed. The ultimate goal will be to develop an instrument to evaluate the preventive psychosocial support system for mental health of children aged 4–10 years across sectors in a community or region of interest.

## 2. Methods/Design

### Study Design

This study, embedded in a German Center for Mental Health (www.dzpg.de) project employs a sequential mixed methods approach, combining the following phases: (1) desk research of existing services, (2) qualitative interviews with professionals from the three sectors – health, education, and social care (H-E-S; see table 1 for definitions), and (3) a quantitative online survey completed by professionals across the three sectors and all institutions of the regional psychosocial support system. Based on these findings, the psychosocial support system for children and their families is mapped in terms of services. To assess collaboration as an integral feature of the psychosocial support system, a social network analysis will be integrated in phase 2 and phase 3. Figure 1 shows the structure of our study with a detailed presentation of the individual phases of data collection and analysis.

**Figure 1.**
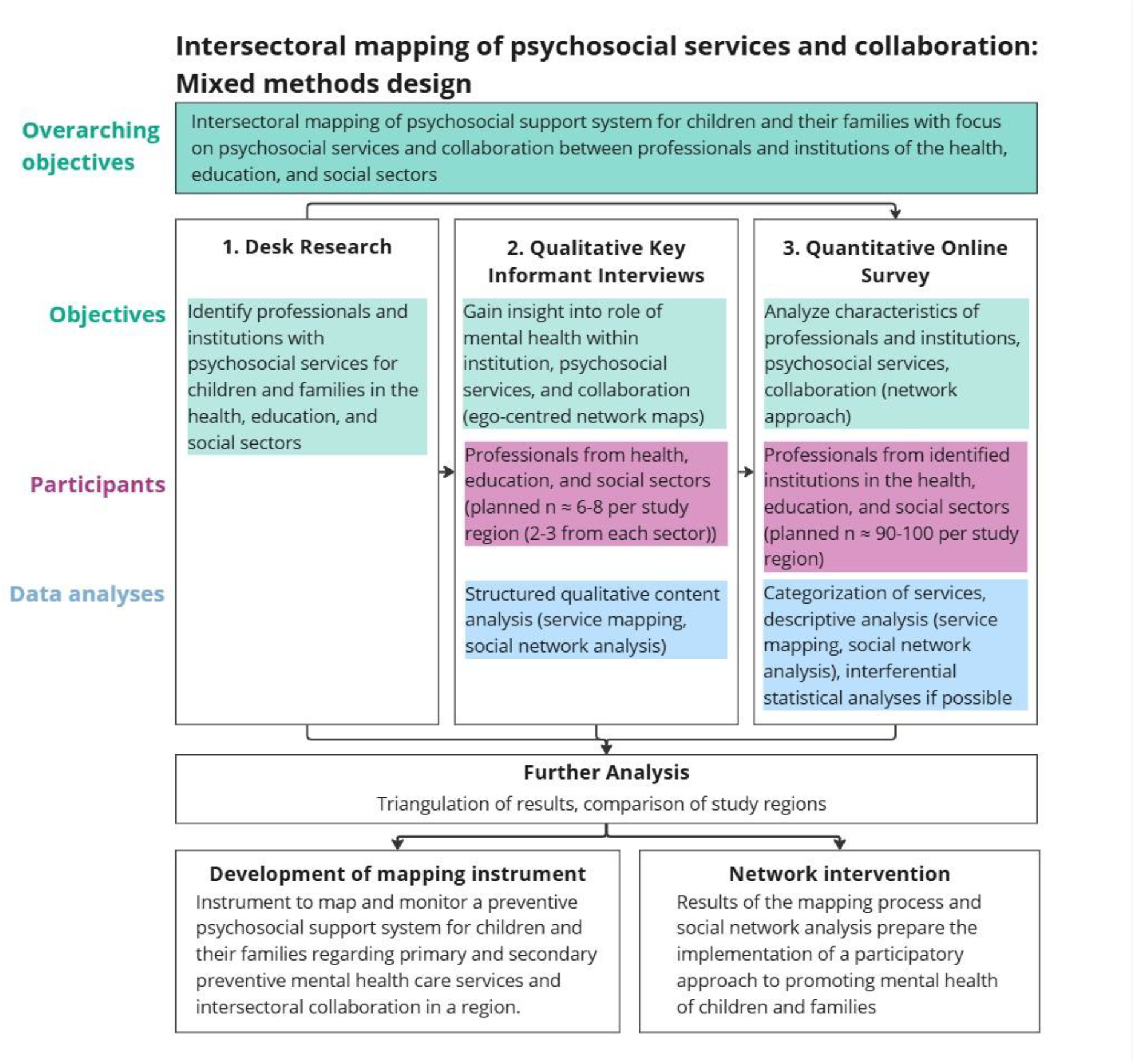
Overview of the study design

### Theoretical underpinnings

The descriptive and analytical approach draws inspiration from systems thinking (15, 16, 38). This perspective involves mapping and analyzing the psychosocial support landscape for children and families as a potentially cohesive system that consists of three different subsystems – health, education, and social sector. Each of these subsystems operates according to its own logic, shaped by variations in financing, governance sponsorship, and underlying motives. These subsystems are further defined by different entities (e.g., institutions) and elements (e.g., service providers, i.e., professionals), which are linked through collaboration and networks. The elements take on roles or functions within the larger system, while its overall output (e.g., supporting children in their psychosocial needs) depends on system dynamics and the patterns of interconnectedness that emerge over time.

This system includes services, service providers, institutions, financial flows, and sponsorship of institutions, as well as collaboration (e.g., referral logics) within and between the H-E-S sectors. The system’s infrastructure and its interconnectedness are depicted by specifically looking at institutions involved, their characteristics (e.g., geographic location, sponsorship), and available psychosocial services for children and their families within the study regions as well as intersectoral collaboration between professionals and institutions in and between the health, education, and social sectors. By mapping this infrastructure, we seek to understand the structure and functioning of the preventive psychosocial support system for children and their families across sector boundaries at community level. At the same time, mapping the psychosocial support system can be used to identify gaps in services and needs-orientation and to identify opportunities for improvement (e.g., indicating areas where to improve intersectoral collaboration). This may provide a foundation for assessing the adequacy of the (regional) psychosocial support system.

Also inspired by the systems perspective, we assume that services pursue certain functions, e.g., providing information, counseling, carrying out diagnostics etc. As part of the mapping of these services, we aim to abstract, systematize and analyze the identified services based on their function(s). We assume that psychosocial services differ in their descriptions or are provided by different professionals but pursue similar or even the same functions. A categorization of services and characterization of service functions (e.g., information, counseling, education) is useful to view psychosocial support from a systems and more generalizable perspective and helps to understand overarching structures in the preventive, psychosocial support system. Furthermore, a conclusion can be made as to which service functions are available, missing or should be available in the system to ensure adequate prevention.

### Definitions

**Table 1.** Definitions.

| Concept | Definition |
| --- | --- |
| Health Sector (H) | The health sector comprises services and institutions that aim to maintain and promote health, including prevention, diagnosis, treatment, and rehabilitation of diseases, as well as health promotion, e.g., hospitals, doctors' surgeries, pharmacies, nursing services, and health authorities (World Health Organization 2024b). |
| Education Sector (E) | The education sector comprises institutions and measures that aim to impart knowledge and skills. This includes early childhood education and care, schools, universities, and vocational schools. The sector is responsible for formal, non-formal, and informal education throughout life (39, 40). |
| Social Sector (S) | The social sector comprises institutions and measures aimed at promoting social justice and integration, as well as social participation. This includes social security, social assistance, child and youth welfare, assistance for the disabled, and counseling services (41, 42). |
| Intersectoral collaboration | There is no definitional consensus on intersectoral collaboration (Hall et al. 2019; World Health Organization 2024a). We are guided by “a broad definition of intersectoral collaboration for mental health as: any planning, information, and resource sharing to institute mental health care between organizations from different sectors (i.e., public, private, not-for-profit) and/or across thematic areas (i.e., health, social care services)” (43). |
| Mental health | Mental health is a state of mental well-being that enables people to cope with the stresses of life, realize their abilities, learn and work well, and contribute to their community. It is an integral component of health and well-being that underpins our individual and collective abilities to make |
|  | decisions, build relationships, and shape the world we live in. Mental health is a basic human right. And it is crucial to personal, community, and socio-economic development. (...) Mental health conditions include mental disorders and psychosocial disabilities, as well as other mental states associated with significant distress, impairment in functioning, or risk of self-harm (44).<br>With respect to children, an emphasis is placed on the developmental aspects, for instance, having a positive sense of identity, the ability to manage thoughts, emotions, as well as to build social relationships, and the aptitude to learn and to acquire an education, ultimately enabling their full active participation in society (7). |
| Mental health disorder | A mental health disorder is characterized by a clinically significant disturbance in an individual's cognition, emotional regulation, or behavior. Mental health disorders can be diagnosed by the International Classification of Diseases 11 (44). |
| Mental health problems | Mental health problems predispose individuals to the development of mental health disorders and, even if thresholds for an explicit mental health disorder are not exceeded (3). They encompass, for example, emotional or behavioral problems such as sadness, aggression, insomnia, difficulties at school, in the family and/or in the social environment (2). |
| Primary prevention | Primary prevention refers to actions aimed at avoiding the manifestation of a disease (this may include actions to improve health through changing the impact of social and economic determinants on health; the provision of information on behavioral and medical health risks, alongside consultation and measures to decrease them at the personal and community level; nutritional and food supplementation; oral and dental hygiene education; and clinical preventive services such as immunization and vaccination of children, adults, and the elderly, as well as vaccination or post-exposure prophylaxis for people exposed to a communicable disease) (45). |
| Secondary prevention | Secondary prevention deals with early detection, which improves the chances for positive health outcomes. This comprises activities such as evidence-based screening programs for early detection of diseases or for prevention of congenital malformations; and preventive drug therapies of proven effectiveness when administered at an early stage of the disease (45). |

### 2.3 Setting

This study will be implemented in two socioeconomically disadvantaged city districts, one from a major city in the west (= study region 1) and one from a major city in the east of Germany (= study region 2). The districts have a population of approximately 20.000 and 45.000 people and are characterized by high poverty rates, a large proportion of migration, and greater remoteness, compared to most other districts in the city. Table 2 shows the characteristics of the city districts.

**Table 2.**
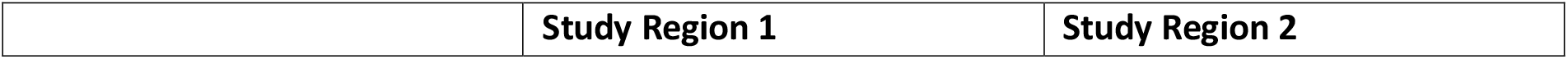
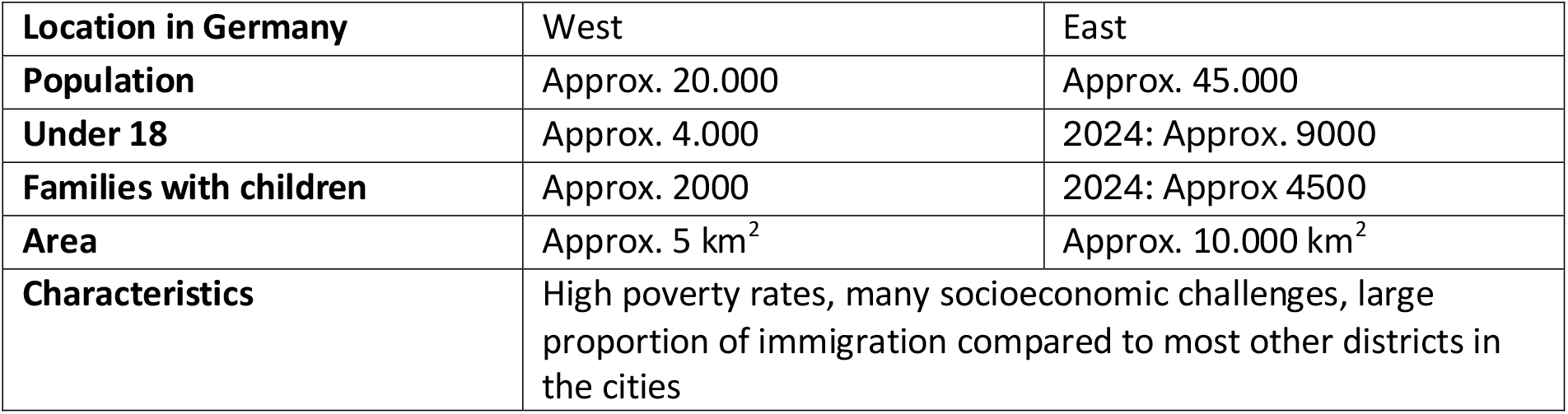
Characteristics of the study regions.

|  | Study Region 1 | Study Region 2 |
| --- | --- | --- |

Table 2. Characteristics of the study regions
| Location in Germany | West | East |
| --- | --- | --- |
| Population | Approx. 20.000 | Approx. 45.000 |
| Under 18 | Approx. 4.000 | 2024: Approx. 9000 |
| Families with children | Approx. 2000 | 2024: Approx 4500 |
| Area | Approx. 5 km<sup>2</sup> | Approx. 10.000 km<sup>2</sup> |
| Characteristics | High poverty rates, many socioeconomic challenges, large proportion of immigration compared to most other districts in the cities |  |

The names of the cities and city districts will remain anonymous in publications.

### 2.4. Study population and recruitment

This study analyzes the psychosocial support system for children and their families at community level, with professionals from institutions serving as the primary source of information. This study will include key professionals from support and care institutions with primary or secondary prevention psychosocial services for children and their families, as well as key professionals working with children in their everyday settings (e.g., schools, early childhood education centers, youth welfare facilities, counseling centers) across the H-E-S sectors. The diversity of sponsorships (public, private) and institutions (e.g., medical practices, schools, counseling centers) will be taken into account. Relevant institutions located outside the study regions, yet providing services to these regions, will also be considered (e.g., pediatric hospitals, public health departments). As not all services focus exclusively on children up to the end of primary school, some of the services surveyed target both children and young people or even whole families.

Through initial desk research and leveraging the study team’s professional networks, institutions and professionals will be identified that fall within the defined study population described earlier. The aim is to compile comprehensive lists of all institutions that potentially form part of the psychosocial support system across the H-E-S in the study regions. From these lists, at least 2-3 key professionals from institutions of each sector will then be contacted for participation in an interview, following a purposeful sampling strategy. Interview participants are the head or deputy management of an institution, or a professional that was recommended by the head of institution. The initial list will be refined throughout the study, incorporating recommendations from the interview participants. For participation in the online survey, all institutions on the lists (including those already participating in the interviews) will be invited. The online survey will be accessed via a QR code or a link sent to the participants by email or distributed by the management within the institution.

### 2.5 Data collection

#### Desk Research

With the help of a desk research, an attempt will be made to compile as comprehensive a list as possible of the services, institutions and contact persons in the psychosocial support system across the H-E-S sectors in the study regions. The lists may be supplemented through information collected during further phases of this study (interviews). Publicly available online information on psychosocial services will be collected and systematically processed according to sector, institution, and geographic location. This investigation will be carried out simultaneously by two research team members for the two study regions, using keyword searches in common online search engines and the study team’s local professional networks. This will involve using generic terms for support services (e.g., preventive mental health services, psychosocial support services, services for families) in combination with the study regions and checking search results for suitability of geographic location, goal of the service and target group.

#### Qualitative Key Informant Interviews

Guideline-based interviews lasting approximately 60 to 90 minutes will be conducted with at least 2-3 key informant professionals from the H-E-S sectors. In total, approximately 6-9 interviews will be conducted for each region. The interview process will be based on a semi-structured guideline in which narrative questions and structuring follow-up questions are combined. The interview guideline will include questions on the type of the professional’s institution and their role within it, on psychosocial services for children and families, as well as questions about the institution’s collaboration, i.e., network partners and the type and frequency of collaboration with them. The interview guide will be pretested with professionals from the H-E-S sectors in other regions before. The interviews will be audio-recorded, transcribed verbatim, and pseudonymized.

#### Ego-centered Network Map Assessment

As part of the key informant interviews, ego-centered network maps (46) will be created, in which professionals enter the relevant institutions and professionals with whom their institution collaborates in the context of psychosocial support for children and their families. These will be prioritized according to importance and placed on the ego-centered network map (see Figure 2). The ego-centered approach of the social network analysis in the key informant interviews will be used to supplement missing services of the desk research accordingly.

**Figure 2.**
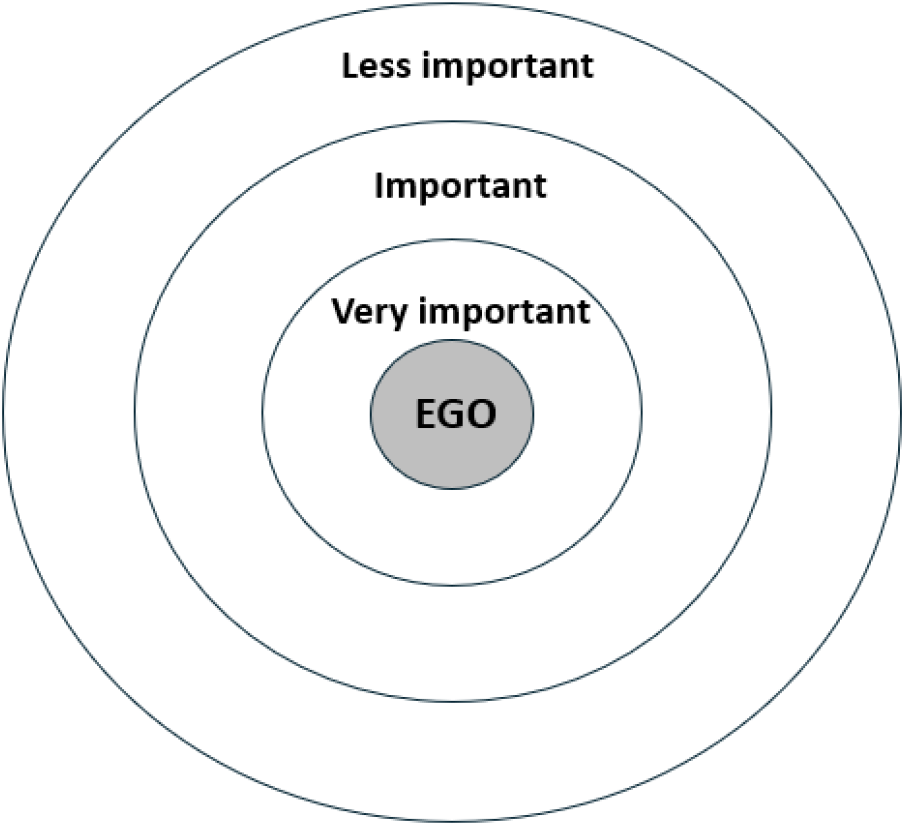
Template of ego-centered network map visualizing collaboration with other professionals/institutions

#### Quantitative Online Survey

An online survey of professionals from institutions in the psychosocial support system will be conducted. As many professionals of the regional support system as possible should take part in this online survey. The number of cases expected depends on the institutions identified in the desk research and the ones participating. The aim is to collect information about the psychosocial services offered by the professionals/institutions, the skills and qualifications of the professionals, and their collaboration with other institutions and professionals.

Professionals will be asked about their institution and their professional background. Furthermore, they will be asked about psychosocial services for children and families including their content, target group, type of services (e.g., outreach service, fixed group service, individual help), service components (e.g., information, counseling, education), psychosocial focus (e.g., behavioral problems, questions about the child’s development and upbringing, family conflicts), funding (e.g., fixed or case-by-case financing), and waiting time. Moreover, professionals will be given a list of other institutions and professionals and will be asked whether they have collaborated with them in the last 12 months regarding case-related psychosocial support for children and families, followed by questions on the type of collaboration (e.g., direct collaboration on a case, referral of children to this network partner), frequency, and satisfaction with the collaboration.

The items of the questionnaire will be inspired by the ESMS and the CAMHS as established mapping instruments but will be adapted to the target group and aim of the study (prevention of mental illness in children) and the community level (27, 29). Questions on collaboration are partly based on established questionnaires from the “Frühe Hilfen” (47).

To increase the comprehensibility of the questions and to identify any problems in the handling of the online survey at an early stage, the online survey will be subjected to a detailed pre-test and adapted if necessary before it is sent out.

#### Ego-centered Network Map Assessment

A network mapping assessment will be implemented into the quantitative online survey. For practical reasons, collaboration with the various professionals and institutions will be surveyed collectively rather than individually at the level of single professionals. For example, we will ask whether participants collaborate with pediatricians in general, instead of listing each pediatric practice individually.

### 2.6 Categorization of services

To apply the systems perspective to identified psychosocial services for children and families, a systematic approach is taken to structure services. For this purpose, a categorization system will be developed that groups the services according to specific aspects, e.g., sector, institution, target group, to create a structured basis for the analysis.

Categories from previous mapping tools, e.g., type/form of service, location, accessibility, provider, availability, target group, funding, will inform the development of the data collection materials and implementation of the study (e.g., interview guidelines, online survey) in order to generate data that enables a basic categorization of the services (27). At the same time, the findings from the individual study phases will potentially lead to additions to the categorization system and the subsequent data collection.

The basic categorization of services according to the three sectors (H-E-S) and the institution will be supplemented by insights gained during the qualitative interviews and quantitative online survey, such as the target group of the service (e.g., children of certain age groups, parents, professionals), the function of the service (e.g., counseling, diagnostics), the psychosocial focus (e.g., behavioral problems, financial difficulties, family conflicts), the funding (e.g., by health insurance, by the authorities), and the waiting time. Other possible categories include the type of prevention (e.g., primary, secondary) and the geographical accessibility. To this end, we will additionally draw on theoretical models, such as those for behavioral change (Behavior Change Wheel) and the Conceptual Framework for Public Mental Health (26, 48).

To interview professionals about the functions of the services they provide, we will draw on theoretical concepts (see above) as well as desk research (phase 1 of the study) and qualitative interviews (phase 2 of the study). The functions developed in such way will then be discussed within the team and piloted with professionals from various professions and institutions. Ultimately, they will be used in the quantitative survey and the categorization system.

### 2.6 Data analysis

#### Desk Research

The study team will review and discuss the results of the desk research. By comparing the results for the two study regions, the study team will reach a consensus on the services to be identified for the preventive psychosocial support of children and their families. Additionally, the study team will agree on how to categorize the identified services by institutions across the H-E-S sectors. This categorization will be incorporated into the quantitative survey which examines collaboration of professionals from the various types of services and sectors as well as into the overall categorization of services derived from the triangulation of the different study phases.

#### Qualitative Data (Key informant interviews, focus groups)

The interviews and focus groups will be coded by two researchers independently using MAXQDA 2024 (VERBI Software, Version 24.4.1, Berlin) and analyzed using qualitative content analysis based on Kuckartz (comparable to the method described by Creswell) (49, 50).

Deductive categories will be defined based on the interview guide and then supplemented by inductive categories during the coding process. Specifically, we will examine the role of children’s mental health within the institution, which preventive psychosocial services are offered, who they are intended for, the format in which they are provided, and the specific activities carried out in these services. We will also examine collaboration with other professionals or institutions, enablers and barriers as well as requirements for collaboration, resulting in an ego-centered network map.

Each interview transcript will be coded by two members of each study team. The coding and results will be discussed within the study team and revised if necessary. The ego-centered network maps will also be coded. For example, the importance of collaboration will be coded in evaluative categories (less important, important, very important) and network partners will be coded according to sector, institution, and/or profession. Transcripts of the interviews and focus groups will also be integrated into the interpretation process.

#### Quantitative Data (Online Survey)

Data from the quantitative survey will be pseudonymized at institutional level, i.e., each institution will be assigned a pseudonym and data will be analyzed at institutional level. The data will be analyzed descriptively using the statistical software SPSS (IBM SPSS Statistics) or R. Open response options will be categorized using the topic modeling procedure in MAXQDA or R or individually and fed back into the dataset. The analyzes will be primarily descriptively comparing the results between the study region, the target groups, and the sectors. If the data allows, inferential statistical analyses will be performed. As part of the analysis of the quantitative data, we will analyze the characteristics of the institutions and the services (e.g., content, target group, form, psychosocial focus, funding) as well as collaboration (i.e., who the institutions work with, in what form, how often and to what extent of satisfaction). Accordingly, we aim to map the overall network of the two study regions.

#### Triangulation of the results of the study phases and comparison of the results from the two study regions

Once this study has been completed, the results of the individual study phases will be analyzed concerning consistency of results and their complementarity via triangulation (51). Triangulation serves the purpose of identifying unique and shared findings across the two study regions and the individual study phases. The aim of consolidating the results from the individual study phases will ultimately be to refine and enhance the overall map of the psychosocial support system, completing the picture as comprehensively as possible. The results from the individual study phases will serve the categorization of preventive psychosocial services that will be developed (see below). The results of the two study settings will be analyzed in relation to each other and compared with regard to available services and collaboration. For example, existing services, institutions, and collaboration can be compared. Any similarities or differences contribute to a more comprehensive understanding of potential phenotypes of ‘systems’ of preventive, psychosocial support. With a view to the two study settings and contextual factors, fundamental differences might be identified and possible causalities determined qualitatively.

#### Patient and public involvement

Patients and public were not involved in the development of this study protocol. The study design was refined by the research team based on scientific literature and expertise. No patient or public involvement is planned in recruitment or data collection; only professionals across the relevant sectors will be surveyed. Patient and public involvement will be considered in future studies based on this mapping study.

## 3. Discussion

To strengthen the prevention of mental health problems in children, the WHO emphasizes the promotion of intersectoral collaboration between professionals (21, 52). To promote intersectoral collaboration between professionals, a better understanding of psychosocial prevention and intersectoral collaboration is necessary (43).

Established mapping approaches to date do not concentrate on community level neither consider psychosocial services in different sectors or preventive services for children (27–30, 32). Therefore, a community mapping approach across the H-E-S sectors with a focus on preventive services for children and families is promising for developing a systematically sound prevention approach for a population in terms of systems thinking (31, 38, 53).

To our knowledge, this is the first attempt at an intersectoral mapping of the psychosocial support landscape for children and their families within and between the three sectors (H-E-S) at community level. The mapping considers institutions, professionals, and services. The focus of this study lies on primary and secondary prevention in the field of mental health for children.

The landscape of psychosocial services, professionals, and institutions initially presents a heterogeneous coexistence of different interventions, institutional logics, and responsibilities. Only by systematically closing existing gaps, through successful, structured collaboration between the institutions and sectors, and by acknowledging patient or client needs involved is it possible to create a functionally coherent system for the prevention of mental illness in children and families from a systems perspective. This would be an inherent part of a client-centered prevention system, given that barriers to seamlessly serve populations and clients in a certain region are still manifold and overcoming those has been shown to improve preventive potential of services (54, 55). Only structure-forming mechanisms such as intersectoral collaboration, feedback loops, shared goals, and coordinated exchange processes acknowledging clients’ needs can transform a fragmented “landscape” into a learning and adaptable preventive system (15, 16, 38).

This mapping will help to better understand the structure and functioning of psychosocial prevention for children and their families, with the goal of promoting mental health from a systems perspective (31, 38). This in-depth understanding might allow identifying gaps in care and developing starting points for improvement of the effectiveness and reach of services. While there are no mapping or monitoring instruments for preventive psychosocial services for children at the age of 4-10 and in different sectors yet (27, 31), this study serves to develop a feasible and at the same time solid approach with a clear methodology and categorization system for the mapping and evaluation of preventive, psychosocial services for children and their families.

### Expected Outcomes and Benefits

Expected results include an overview of existing regional psychosocial services for children and their families, with professionals, institutions, and services involved. A categorization of services can provide information about the structure and availability of preventive services as well as priorities within the sectors and the community as a whole. If a sufficient number of professionals participate in this study, a comprehensive overview of local preventive psychosocial services and collaboration in the study regions will be possible. This could help local families find suitable support services and local professionals to better collaborate and establish seamless referral processes for individualized client profiles.

Analyzing collaboration will help determine the actual collaboration in the respective system and the perspectives of the professionals on collaboration. By linking the mapping of psychosocial services with the results of the network analysis, it might be possible to determine in which areas preventive psychosocial service provision is particularly strong or weak (56). Also, care gaps and needs for services can be identified.

Mapping and understanding the psychosocial support system might be important for initiating structural changes, with the objective of promoting the mental health of children more efficiently and with higher reach. For example, the mapping, but also the network analysis, could serve as a starting point to a tailored intervention to support collaboration. Building on insights from the social network analysis, use cases can be developed to illustrate typical scenarios and challenges within the psychosocial support system. These can guide structured discussions with professionals to clarify roles, responsibilities, and referral pathways. The qualitative analysis of the factors that promote and hinder intersectoral collaboration could also provide important insights into potential for the development of more effective preventive system and collaboration structures.

### Limitations and challenges

The intersectoral analysis of psychosocial services and collaboration is associated with several challenges in its implementation.

Although we consider the complexity and include a wide range of sectors with their variety of primary and secondary prevention services for different target groups (children, families), our approach may not be differentiated enough to truly capture all the implications of this complexity. As the structures and tasks of the various professionals vary strongly in and between the health, education, and social sectors, it may not always be possible to ensure that the survey instruments fully reflect the specific situation of each service.

Another limiting factor is, that only professionals and not clients are surveyed. Therefore, our study does not provide any information on the evaluation and use of the support system by the intended client group itself. However, future surveys at the client level of psychosocial services could be used to compare the results of the professionals with the results of the clients. It is important to note, however, that through its mapping approach, we can only offer a snapshot of a time-dynamic complex ‘system.’

Last, the study is based on the voluntary participation of the institutions and their professionals. Therefore, participants might have an intrinsic motivation, which could lead to selection or non-response biases (57); on the other hand, this could mean that not all institutions participate and therefore the support system cannot be captured completely.

### Outlook

As there is no blueprint for the intersectoral mapping of psychosocial services and collaboration for children and families, we plan to use the results of the study and the experience gained to develop an instrument that can be used in future for mapping and monitoring the characteristics and the form of the psychosocial support system for children and their families in a defined region. As this is a very complex procedure, it should consist of individual modules that can be adapted to specific age groups or different administrative levels (e.g., municipalities, federal states). The results of the study and the knowledge gained about the process of data collection and data analysis will be incorporated into the concrete application form of the mapping instrument. The mapping instrument will describe the methodological steps, analysis strategies, and categorization of psychosocial services. Such a categorization can, for example, provide information on which services already exist and where gaps exist in the prevention system or in which areas collaboration must be boosted. In this line, the instrument could be used at community level for further developing existing collaboration and services for the mental health of children. To enable service mapping in other regions in the future and to update it regularly and efficiently in response to potential changes in the service landscape, AI-based automated searches as part of desk research are an option. The results of the mapping could inform such automated searches, for example, by providing search terms.

## Data Availability

No data is included in the manuscript.

## 4. List of abbreviations

DZPG: German Center for Mental Health (Deutsches Zentrum für Psychische Gesundheit)
WHO: World Health Organization
H-E-S: Health, education, and social sectors

## 5. Declarations

The Ethics Committee of the Faculty of Psychology at the Ruhr University Bochum has issued a favorable ethics vote (ID Nr: 876, 13/12/2023).

All authors have approved the manuscript for submission.

-Availability of data and materials – Not applicable

## Author statement

A.R. and S.B. conceptualization, writing of the original draft, creation of figures, revision of the manuscript based on feedback from the other co-authors

F.D.B. conceptualization, development of methods, revision of manuscript, provision of critical feedback, supervision

B.R., S.W., S.K., and A.A. conceptualization, revision of manuscript, provision of critical feedback, supervision

L.P., B.V., S.S. revision of manuscript, provision of critical feedback

## Footnotes

1 In the following, we will use the short term psychosocial services

## References

1. Sevecke K, Wenter A, Haid-Stecher N, Fuchs M, Böge I. Die psychische Gesundheit unserer Kinder und Jugendlichen und deren Behandlungsmöglichkeiten im Drei-Länder-Vergleich (Ö, D, CH) unter Berücksichtigung der Veränderungen durch die COVID-19-Pandemie. Neuropsychiatr 2022; 36(4):192–201.

2. Hölling H, Schlack R, Petermann F, Ravens-Sieberer U, Mauz E. Psychische Auffälligkeiten und psychosoziale Beeinträchtigungen bei Kindern und Jugendlichen im Alter von 3 bis 17 Jahren in Deutschland - Prävalenz und zeitliche Trends zu 2 Erhebungszeitpunkten (2003-2006 und 2009-2012): Ergebnisse der KiGGS-Studie - Erste Folgebefragung (KiGGS Welle 1). Bundesgesundheitsblatt Gesundheitsforschung Gesundheitsschutz 2014; 57(7):807–19.

3. RKI. Psychische Gesundheit in Deutschland. Erkennen - Bewerten - Handeln: Schwerpunktbericht Teil 2 - Kindes-und Jugendalter; Fokus: Psychische Auffälligkeiten gemäß psychopathologischem Screening und Aufmerksamkeitsdefizit-/Hyperaktivitätsstörung (ADHS). Berlin; 2021.

4. Lorenz S, Ulrich SM, Sann A, Liel C. Self-Reported Psychosocial Stress in Parents With Small Children. Dtsch Arztebl Int 2020; 117(42):709–16.

5. Eickhorst A, Liel C. Design und Methoden der Studienfolge “Kinder in Deutschland - KiD 0-3”: Faktenblatt 1 zur Prävalenz-und Versorgungsforschung der Bundesinitiative Frühe Hilfen. Köln; 2020.

6. Eickhorst A, Schreier A, Brand C, Lang K, Liel C, Renner I et al. Inanspruchnahme von Angeboten der Frühen Hilfen und darüber hinaus durch psychosozial belastete Eltern. Bundesgesundheitsblatt Gesundheitsforschung Gesundheitsschutz 2016; 59(10):1271–80.

7. World Health Organization. Comprehensive Mental Health Action Plan: 2013–2030. Geneva; 2021. Available from: URL: https://iris.who.int/bitstream/handle/10665/345301/9789240031029-eng.pdf?sequence=1.

8. Kessler RC, McLaughlin KA, Green JG, Gruber MJ, Sampson NA, Zaslavsky AM et al. Childhood adversities and adult psychopathology in the WHO World Mental Health Surveys. Br J Psychiatry 2010; 197(5):378–85.

9. Klasen F, Meyrose A-K, Otto C, Ravens-Sieberer U. Psychische Auffälligkeiten von Kindern und Jugendlichen in Deutschland. Monatsschrift Kinderheilkunde 2017; 165(5):402–7.

10. Pössel P, Hautmann C. Prävention psychischer Störungen im Kindes-und Jugendalter. In: Döpfner M, Hautzinger M, Linden M, editors. Verhaltenstherapiemanual: Kinder und Jugendliche. Berlin, Heidelberg: Springer Berlin Heidelberg; 2020. p. 165–9 (Psychotherapie: Praxis).

11. Ewert B. Promoting health in schools: Theoretical reflections on the settings approach versus nudge tactics. Soc Theory Health 2017; 15(4):430–47.

12. Herbert B, Strauß A, Mayer A, Duvinage K, Mitschek C, Koletzko B. Implementation process and acceptance of a setting based prevention programme to promote healthy lifestyle in preschool children. Health Education Journal 2013; 72(3):363–72.

13. King L. The settings approach to achieving better health for children. NSW Public Health Bull. 1998; 9(11):128.

14. World Health Organization. Guidelines on mental health promotive and preventive interventions for adolescents: Helping adolescents thrive. Geneva: World Health Organization; 2020. Available from: URL: https://www.ncbi.nlm.nih.gov/books/NBK565375/.

15. Luna Pinzon A, Stronks K, Dijkstra C, Renders C, Altenburg T, Hertog K den et al. The ENCOMPASS framework: a practical guide for the evaluation of public health programmes in complex adaptive systems. Int J Behav Nutr Phys Act 2022; 19(1):33.

16. Traeber-Burdin S, Varga M. How does Systems Thinking support the Understanding of Complex Situations? In: 2022 IEEE International Symposium on Systems Engineering (ISSE). IEEE; 2022. p. 1–7.

17. Roberts JH, Crosland A, Fulton J. “I think this is maybe our Achilles heel…” exploring GPs’ responses to young people presenting with emotional distress in general practice: a qualitative study. BMJ Open 2013; 3(9):e002927.

18. Franzke A, Schultz A. Präventionsangebote - Was beeinflusst die Inanspruchnahme?: Theorie und Methode der Familienbefragung “Kein Kind zurücklassen!”; 2015.

19. Kourgiantakis T, Markoulakis R, Lee E, Hussain A, Lau C, Ashcroft R et al. Access to mental health and addiction services for youth and their families in Ontario: perspectives of parents, youth, and service providers. Int J Ment Health Syst 2023; 17(1).

20. Ekornes S. Teacher Perspectives on Their Role and the Challenges of Inter-professional Collaboration in Mental Health Promotion. School Mental Health 2015; 7(3):193–211.

21. Continuity and coordination of care: a practice brief to support implementation of the WHO Framework on integrated people-centred health services; 2018. Available from: URL: https://integratedcarefoundation.org/wp-content/uploads/2018/08/WHO-practice-brief-on-continuity-and-coordination-of-care.pdf.

22. Jeindl R, Hofer V, Bachmann C, Zechmeister-Koss I. Optimising child and adolescent mental health care - a scoping review of international best-practice strategies and service models. Child Adolesc Psychiatry Ment Health 2023; 17(1):135.

23. Babatunde GB, van Rensburg AJ, Bhana A, Petersen I. Identifying multilevel and multisectoral strategies to develop a Theory of Change for improving child and adolescent mental health services in a case-study district in South Africa. Child Adolesc Psychiatry Ment Health 2022; 16(1):45.

24. Metzner G, Horstmann S, Barth M, Giesler JM, Jünemann S, Kaier K et al. Evaluation of a cross-sectoral care intervention for families with psychosocial burden: a study protocol of a controlled trial. BMC Health Serv Res 2022; 22(1):475.

25. Campion J. Public mental health: key challenges and opportunities. BJPsych Int 2018; 15(3):51–4.

26. Wahlbeck K. Public mental health: the time is ripe for translation of evidence into practice. World Psychiatry 2015; 14(1):36–42.

27. Johnson S, Kuhlmann R. The European Service Mapping Schedule (ESMS): development of an instrumentfor the description and classificationof mental health services. Acta Psychiatr Scand 2000; 102(405):14–23.

28. Romero-López-Alberca C, Gutiérrez-Colosía MR, Salinas-Pérez JA, Almeda N, Furst M, Johnson S et al. Standardised description of health and social care: A systematic review of use of the ESMS/DESDE (European Service Mapping Schedule/Description and Evaluation of Services and DirectoriEs). Eur Psychiatry 2019; 61:97–110.

29. Signorini G, Singh SP, Boricevic-Marsanic V, Dieleman G, Dodig-Ćurković K, Franic T et al. Architecture and functioning of child and adolescent mental health services: a 28-country survey in Europe. Lancet Psychiatry 2017; 4(9):715–24.

30. Salvador-Carulla L, Amaddeo F, Gutiérrez-Colosía MR, Salazzari D, Gonzalez-Caballero JL, Montagni I et al. Developing a tool for mapping adult mental health care provision in Europe: the REMAST research protocol and its contribution to better integrated care. Int J Integr Care 2015; 15:e042.

31. Stansfield J, Cavill N, Marshall L, Robson C, Rutter H. Using complex systems mapping to build a strategic public health response to mental health in England. JPMH 2021; 20(4):286–97.

32. Kilicel D, Crescenzo F de, Barbe R, Edan A, Curtis L, Singh S et al. Mapping Child and Adolescent Mental Health Services and the Interface During Transition to Adult Services in Six Swiss Cantons. Front Psychiatry 2022; 13:814147.

33. Küster E-U, Peterle C. Netzwerkkoordinierende in den Frühen Hilfen: Faktenblatt zu den NZFH-Kommunalbefragungen. Köln; 2023.

34. Werkbuch Präventionskette: Herausforderungen und Chancen beim Aufbau von Präventionsketten in Kommunen; 2013. Available from: URL: https://www.praeventionsketten-nds.de/fileadmin/media/downloads/Werkbuch-Praeventionskette_Doppelseite.pdf.

35. Bock F de, Dietrich M, Rehfuess E. Evidenzbasierte Prävention und Gesundheitsförderung. Memorandum der Bundeszentrale für gesundheitliche Aufklärung (BZgA); 2021.

36. Evans RE, Moore G, Movsisyan A, Rehfuess E. How can we adapt complex population health interventions for new contexts? Progressing debates and research priorities. J Epidemiol Community Health 2021; 75(1):40–5.

37. Hawe P, Shiell A, Riley T. Complex interventions: how “out of control” can a randomised controlled trial be? BMJ 2004; 328(7455):1561–3.

38. Meadows DH. Thinking in systems: A primer. [Nachdr.]. White River Junction, Vt: Chelsea Green Pub; 2011. Available from: URL: http://www.loc.gov/catdir/enhancements/fy0905/2008035211-b.html.

39. OECD. Education at a Glance 2023. OECD; 2023.

40. UNESCO. Global Education Monitoring Report 2020: Inclusion and education: All means all. Paris. UNESCO; 2020.

41. International Labour Organization. World Social Protection Report 2020-22: Social Protection at the Crossroads - in Pursuit of a Better Future. Genève 22; 2021. Available from: URL: https://ebookcentral.proquest.com/lib/kxp/detail.action?docID=6941151.

42. United Nations. World Social Report 2023. United Nations; 2023.

43. Hall T, Kakuma R, Palmer L, Minas H, Martins J, Armstrong G. Intersectoral collaboration for people-centred mental health care in Timor-Leste: a mixed-methods study using qualitative and social network analysis. Int J Ment Health Syst 2019; 13:72.

44. Mental health; 2022. Available from: URL: https://www.who.int/news-room/fact-sheets/detail/mental-health-strengthening-our-response.

45. Regional Office for the Eastern Mediterranean. Health promotion and disease prevention through population-based interventions, including action to address social determinants and health inequity; Available from: URL: https://www.emro.who.int/about-who/public-health-functions/health-promotion-disease-prevention.html.

46. Hollstein B, Pfeffer J. Netzwerkkarten als Instrument zur Erhebung egozentrierter Netzwerke; 2010. Available from: URL: http://www.pfeffer.at/egonet/Hollstein%20Pfeffer.pdf.

47. Sann A. Teiluntersuchung: Kooperationsformen. 1. Aufl., 10.10.10. Köln: Nat. Zentrum Frühe Hilfen; 2010. (Materialien zu Frühen Hilfen Bestandsaufnahme; vol 2).

48. Michie S, van Stralen MM, West R. The behaviour change wheel: a new method for characterising and designing behaviour change interventions. Implement Sci 2011; 6:42.

49. Kuckartz U. Qualitative Inhaltsanalyse: Methoden, Praxis, Computerunterstützung. Weinheim, Basel: Beltz Juventa; 2012.

50. Creswell JW. Educational research: Planning, conducting, and evaluating quantitative and qualitative research. 4th. ed. Boston: Pearson.

51. Kuckartz U. Mixed-Methods-Datenanalyse. In: Kuckartz U, editor. Mixed Methods: Methodologie, Forschungsdesigns und Analyseverfahren. Wiesbaden: Springer VS; 2014. p. 99–122 (Lehrbuch).

52. World Health Organization. Multisectoral action for mental health; 2019. Available from: URL: https://iris.who.int/bitstream/handle/10665/346538/WHO-EURO-2019-3579-43338-60799-eng.pdf?sequence=1.

53. Popp L, Voigt B, Julich A, König H-H, Brettschneider C, Kulisch LK et al. Urban Mental Health: Förderung der psychischen Gesundheit von jungen Menschen in der Stadt. Kindheit und Entwicklung 2025; 34(1):23–32.

54. Slack KS, Berger LM, Reilly A, Reynders R, Cai JY. Preventing child protective services system involvement by asking families what they need: Findings from a multi-site RCT of the community response program (CRP). Child Youth Serv Rev 2022; 141:106569.

55. Reinhart A, Alayli A, Beierle S, Löffler A, Reißig B, Walper S et al. Barriers to accessing and using preventive mental health services for psychosocially strained children and families in Germany: Perspectives of professionals from different sectors. Prev Med 2025; 200:108392.

56. Luke DA, Harris JK. Network analysis in public health: history, methods, and applications. Annu Rev Public Health 2007; 28:69–93.

57. Liedl B, Steiber N. Führen Online-Befragungen zu anderen Ergebnissen als persönliche Interviews? Eine Schätzung von Moduseffekten am Beispiel eines Mixed-Mode Surveys. Österreich Z Soziol 2024; 49(1):1–22.

